# Benchmarking a massively parallel nucleic acid hybridization platform for monitoring biomarkers of public health significance in wastewater

**DOI:** 10.1101/2025.04.24.25326374

**Authors:** Ocean Thakali, Walaa Eid, Julia Brasset-Gorny, Sean Stephenson, Elizabeth Mercier, Robert Delatolla, Tyson E. Graber

**Affiliations:** Children’s Hospital of Eastern Ontario Research Institute, Ottawa, Ontario K1H 8L1, Canada; Department of Civil Engineering, University of Ottawa, Ottawa, Ontario K1N 6N5, Canada

**Keywords:** Antimicrobial resistance, crAssphage, Nanostring nCounter, direct detection, viruses, wastewater-based epidemiology, wastewater and environmental surveillance

## Abstract

Nucleic acid amplification tests (NAATs) are exquisitely sensitive and specific, able to accurately and quickly monitor vanishingly small amounts of RNA or DNA in wastewater-based public health surveillance applications. Yet, multiplexing samples and target assays is difficult and the dependence of NAATs on enzymes increases false negative rates with inhibitor-rich environmental samples. The Nanostring nCounter system (NNS) is a nucleic acid hybridization platform with a capability for high multiplexing (upwards of 800 probes) that directly counts RNA or DNA biomarkers without reverse transcription or amplification. This study determines the feasibility of direct detection and quantification of genetic markers of public health significance in wastewater samples using NNS and compares the performance with gold-standard NAAT assays targeting antimicrobial resistance (AMR) DNA loci, 16S DNA, SARS-CoV-2 RNA, and the fecal content markers, Pepper mild mottle virus (PMMoV) RNA and crAssphage DNA. We demonstrate that the direct detection and quantification of high- and medium-copy markers, including PMMoV RNA, beta-lactamase, and carbapenemase DNA, in wastewater extracts is both feasible and accurate when benchmarked against gold-standard NAATs. Low-copy detection and monitoring daily trends of SARS-CoV-2 N RNA was achievable but was not as robust as NAATs. Due to the lower analytical sensitivity of NNS compared to NAATs and the requirement of a 18h hybridization step, NNS may not be suitable to provide early warning of incident cases to public health. However, as the scope of wastewater surveillance expands to monitor a broad range of nucleic acid-based biomarkers, the target and sample multiplexing capability of NNS, combined with reduced hands-on time and ease-of-analysis, are distinct advantages. Thus, NNS has a role to play in wastewater and environmental monitoring applications, especially for AMR surveillance.

**Highlights:**

- Nanostring nCounter system (NNS) benchmarked in wastewater surveillance (WWS).
- Markers of antimicrobial resistance, SARS-CoV-2, and feces directly measured by NNS.
- Strong concordance between NNS and nucleic acid amplification tests (NAATs).
- NNS has lower sensitivity compared to NAATs.
- Cost-effective, scalable multiplexing of NNS is an advantage in WWS.

## 1. Introduction

Wastewater surveillance (WWS) has become a valuable tool in recent years, complementing clinical surveillance by monitoring the circulation of infectious disease pathogens and antimicrobial resistant genes (ARGs) that contribute to antimicrobial resistance (AMR) within communities. WWS primarily involves quantification of pathogens over time to reflect the trend of infection in a given wastewater treatment plant catchment. Nucleic Acid Amplification tests (NAATs) such as reverse transcription quantitative PCR (RT-qPCR) or qPCR for RNA or DNA targets, respectively, are the most widely used in WWS to amplify and quantify ARG or pathogen RNA/DNA biomarkers because of low operational cost, quick turnaround time, and very high sensitivity and specificity. However, the common occurrence of PCR inhibitors in wastewater, amplification bias, and variation in quality of reference materials used to construct calibration curves can result in inaccurate or unreliable quantification of target amplicons using NAATs (Hamza and Leifels 2024, Linzner et al., 2024, D’Aoust et al., 2021, Graham et al., 2021, Gonzalez et al., 2020, Casado-Martín et al., 2025). To overcome these challenges, digital PCR (dPCR), including RT-dPCR for RNA targets and its earlier iteration digital droplet PCR (ddPCR), is increasingly employed to quantify nucleic acid biomarkers in environmental matrices (Tiwari et al., 2022). dPCR and ddPCR separate the sample into thousands of individual partitions and provide absolute quantification of the target following enzyme-mediated amplification based on Poisson statistics. These can provide accurate and precise quantification over a dynamic range of up to four orders of magnitude.

Despite the above-mentioned advantages, dPCR is currently more expensive and has low throughput; with target multiplexing requiring substantial assay optimization. Therefore, alternative platforms such as High-throughput qPCR (HT-qPCR) systems (Malla et al., 2022, Ishii et al., 2014), ATOPlex sequencing (Ahmed et al., 2022, Ni et al., 2021), GeneXpert systems (Cepheid, CA, USA) (Daigle et al., 2022, Asadi et al., 2023), DNA microarray (Lee et al., 2006), and Loop-mediated isothermal amplification (LAMP) (Amoah et al., 2021, Bivins et al., 2021) has also been used for wastewater surveillance. All the above-mentioned platforms have their own advantages and disadvantages. For example, a HT-qPCR platform such as the Biomark HD system (Fluidigm, CA, USA) is a good alternative when multiplexing is required, but can require extensive hands-on time for specific target amplification that can also promote cross-contamination. GeneXpert system and LAMP can be viable options for isolated remote communities without established testing laboratories, but these platforms are prone to low analytical sensitivity and are intended to only provide information on the presence or absence of pathogens (Bivins et al., 2021).

The Nanostring nCounter system (NNS) is a platform that relies on massively parallelized nucleic acid-based probe sequences tagged with fluorescent barcodes to directly detect and quantify up to 800 DNA or RNA targets within one sample without the need for enzyme-mediated reverse transcription of RNA, or DNA amplification (Geiss et al., 2008). Sensitivity and specificity are achieved with pairs of biotin-conjugated oligonucleotide capture probes and fluorescently barcoded (up to six fluorophores) oligonucleotide reporter probes that together bind to a ∼100 nt region on the intended target DNA or RNA molecules in-solution during a ∼16-hour incubation (**Fig. 1**). The biotinylated probe-target hybridized molecules are then immobilized to a streptavidin-impregnated imaging surface imaging within a microfluidic chip, non-bound material is washed away, an electric field is applied to ensure correct barcode orientation, and fields of bound molecules are imaged with an automated, high-resolution CCD camera. The labelled barcodes are directly counted and correlate with the concentration of target DNA or RNA. The key advantages of NNS includes: 1) High target multiplexing capability (in principle, 800 probes can be combined/plexed in a single sample run; 2) Up to 12 samples can be run on a single chip, and additional sample plexing can be achieved within a chip lane by using different barcodes; 3) Reverse transcription or DNA polymerase-mediated amplification is not required (although it could be applied to increase sensitivity) and is thus unlikely to be affected by PCR inhibitors; 3) Direct, absolute quantification of target DNA/RNA; 4) A closed, automated system results in less hands-on time (15 minutes) and reduced chances of contamination; 5) Robust quality control with internal controls and straight-forward analysis that does not require reference standards; 6) A system that does not require significant expertise to operate, troubleshoot, or analyze the data and whose assays are easy to build, validate, and add to existing probesets.

**Figure 1.**
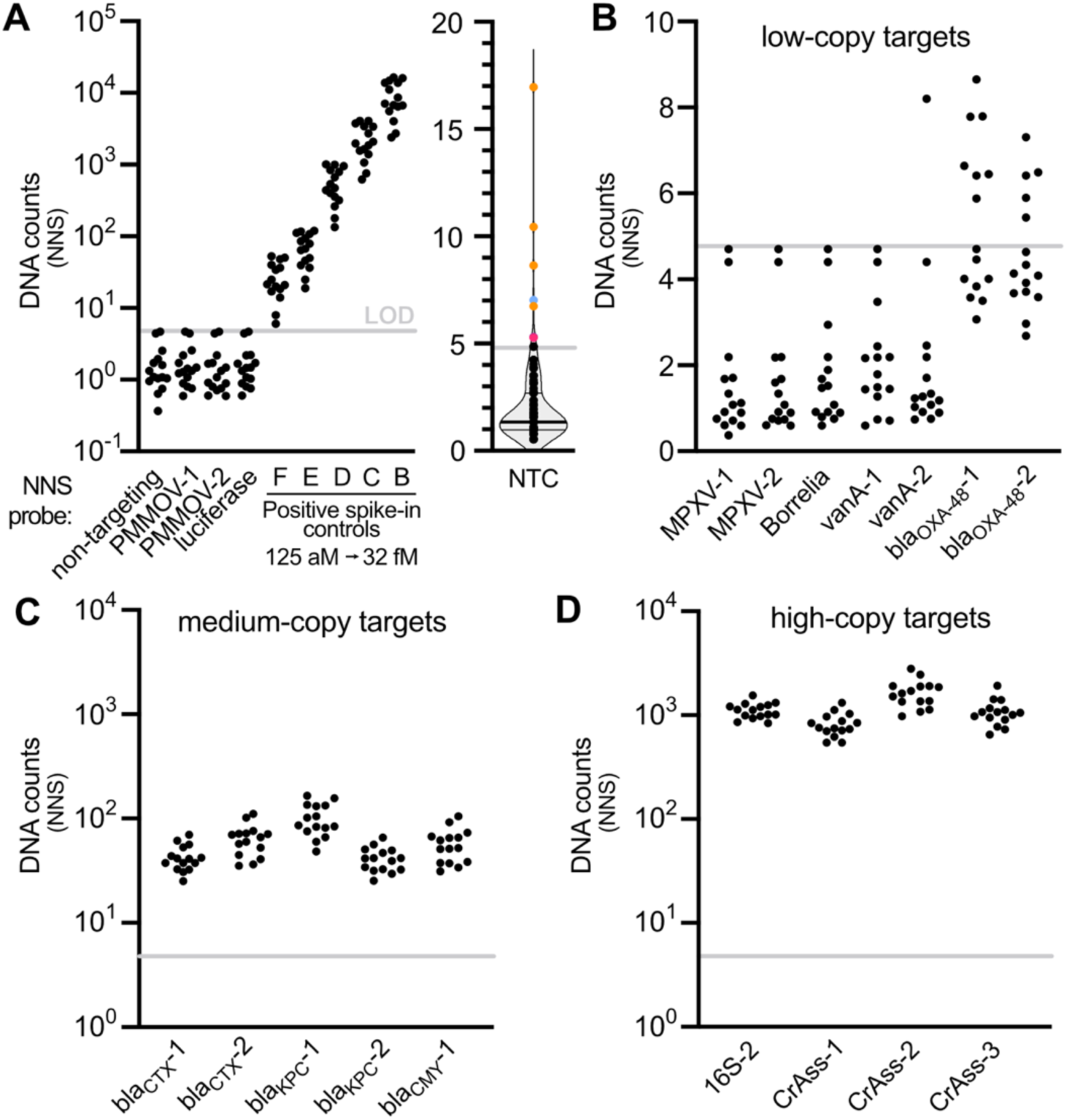
Targets measured using NNS in DNA extracted from primary sludge. (**A**) NNS demonstrates minimal off-target binding of probes. Probe-specific DNA target counts are shown from 5 different sludge-derived DNA extracts and 3 different runs of the same extract. Probes that are non-targeting (ERCC probes), those that target PMMoV (an RNA template that shouldn’t be present in DNA extracts), or *Photinus pyralis* luciferase gene (a DNA template not expected to be present in wastewater extracts) exhibited background counts below the LOD (light grey line). Note “-1” and “-2” denoting two different probes for the same target gene or organism. Distribution of counts for each probe in NTC lanes. 16S (orange) exhibited consistent, background signal close to LOD (rightmost plot). CrAssphage (blue) and vanA (red) exhibited signal above LOD in only one of the three runs and only one probeset and were thus interpreted as stochastic in origin. (**B**) Counts of low-copy targets namely MPXV, *Borrelia spp.* and less prevalent ARGs. (**C**) Counts of medium-copy targets; 16S rRNA genes (multiple species), Crassphages. (**D**) Counts of high-copy ARGs.

Despite the clear potential advantages, only one study has used NNS to quantify targets of interest in environmental samples. Tolman et al quantified DNA biomarkers involved in nitrogen cycle pathway in environmental DNA samples recovered from seawater (Tolman et al., 2024). Past applications of the NNS have been limited to gene expression studies in the fields of immunology, oncology, microbiology, and neuroscience (Chilimoniuk et al., 2024, Eastel et al., 2019, Tekletsion et al., 2018) as well as bacterial and fungal RNA counts in clinical samples (Barczak et al., 2012, Bispo et al., 2023, Grahl et al., 2018).

Based on this background, this study aimed to detect and quantify biomarkers of public health significance in wastewater samples using NNS and compare the performance with digital-droplet PCR (ddPCR) and qPCR. We successfully quantified several AMR DNA biomarkers as well as fecal content RNA biomarkers with NNS from nucleic acid-extracted wastewater samples. Moreover, we were able to directly quantify SARS-CoV-2 RNA that correlated with retrospectively collected gold-standard RT-qPCR signal in primary sludge samples over a 6-week period characterized by a waning COVID-19 outbreak in the community. The findings of this study provide the research community with a starting toolbox of validated nanostring probesets for applying in wastewater surveillance settings and demonstrate the feasibility and utility of this platform, especially with respect to monitoring AMR biomarkers. The platform’s high plexibility allows for rapid, sensitive and quantitative trend monitoring of hundreds of targets with a single sample in a single tube and at a per-sample and instrument cost that falls between qPCR and next-generation sequencing technologies.

## 2. Materials and Methods

### 2.1. Nucleic acid targets

Probes targeting RNA and DNA biomarkers were chosen based on those currently used in many WWS programmes (namely the PMMoV genome and SARS-CoV-2 N RNA, crAssphage DNA, MPXV DNA). Intended as a positive control, we designed probes targeting bacterial 16S rRNA genes which mapped to *E. coli* bacteria amongst others, and that would be expected to be in high concentration in wastewater samples. A probe targeting firefly luciferase RNA served as a negative control, and if the samples were spiked with the targeted RNA could also be used as an extraction recovery control. A probe targeting the 16S rRNA gene of *Borrelia spp.* spirochete bacteria was also included. Several of these species are the causative agents of Lyme disease in humans, contracted via infected ticks, and considered an emergent infectious disease of public health concern in the Ottawa region, normally monitored by a dedicated tick surveillance program. Finally, ARG markers were selected based on their clinical significance and prevalence in wastewater, as determined by qPCR data (Daigle, 2024). Consultations with Jade Daigle and Chand Mangat at the Public Health Agency of Canada were critically important in this regard. Briefly, the *vanA* gene conferring vancomycin resistance, bla OXA-48 gene (bla_OXA-48_; confers carbapenem resistance), bla CTX gene (bla_CTX_; confers resistance to a broad range of β-lactam antibiotics, but not carbapenems), bla KPC (bla_KPC_; confers carbapenem resistance), bla CMY gene (bla_CMY_; confers cephalosporin resistance).

The assays also control probes provided by Nanostring Technologies. These include six synthetic positive control oligonucleotides with concentrations spanning 3 orders of magnitude (0.125-128 fM) along with their matched probesets, together with six negative control probesets that should not bind to the DNA or RNA of any known organism (i.e., non-targeting). These synthetic oligonucleotides and the control probesets are spiked into samples and linear regression of the expected concentration plotted against counts of those positive controls can be used to measure the efficiency of the hybridization and imaging procedure, while the negative controls can help rule out false positives and/or normalize counts against variation of assay inputs.

### 2.2. Concentration of primary sludge samples and extraction of nucleic acids

Primary sludge samples representing 24-hour composited material were collected from a WWTP in Ottawa, Canada and centrifuged at 15000 ×*g* for 10 min at 4 °C to pellet wastewater solids as previously described (D’Aoust et al., 2021). Approximately 100 mg of the pellet was used to extract 100 μL of RNA only using the RNeasy Powerfecal Pro Kit (QIAGEN, Halden, Germany) as per the manufacturer’s instructions. Similarly, ∼250 mg of the pellet was processed to extract 50 μL of DNA only, using the AllPrep PowerViral DNA/RNA kit (QIAGEN) with few modifications. Bead-beating tubes were homogenized using an Omni bead ruptor 24 (Omni International Inc., GA, USA) and prior to the addition of Solution PM5, 50 μL of RNase (concentration:1 ng/μL) was added to the spin column and left at room temperature for 15 min to remove all RNA. The concentration and quality of nucleic acid was then measured with a Nanodrop 1000 spectrophotometer (Thermo Fisher Scientific Inc., DE, USA).

### 2.3. In vitro transcription of SARS CoV-2 N RNA

A plasmid containing the full-length 1.3 kb SARS-CoV-2 N open reading frame (ORF; Wuhan variant) was created using an overlap extension PCR strategy with four synthesized DNA fragments (Thermofisher GeneArt Strings, Germany) that included BamHI and NotI restriction sites in the PCR primers. A BamHI-NotI-digested fragment was then directionally inserted into a pcDNA3.1 vector that contained T7 RNA polymerase start and stop sites by T4 ligation. Clones were selected, DNA extracted, and the plasmid (pcDNA3.SARS-CoV-2 N) was sequence-verified by Sanger sequencing. Full-length N ORF RNA was in vitro transcribed (IVT) using MEGAscript® T7 RNA polymerase, reaction buffer and ribonucleotides (Thermo Fisher Scientific) in a 20 µL reaction with 1 μg of NotI-linearized N plasmid DNA. The reaction was incubated at 37°C for four hours. 1 µL TURBO DNase (Thermo Fisher Scientific) was added and incubated at 37°C for 15 minutes to remove template DNA. IVT RNA was purified using the MEGAclear™ Kit (Thermo Fisher Scientific) per the manufacturer’s protocol and RNA was eluted from the column by incubating with 50 µL Elution Solution at 65–70°C for 10 minutes, followed by centrifugation at room temperature. The N RNA was stored at -80 °C. N copies were determined in diluted IVT reactions by RT-ddPCR using the N1 assay as described below. After the determination of N1-copy number by RT-ddPCR, titration curves of IVT N RNA (∼200-3000 N1-equivalent copies) were subjected to analysis by NNS to validate the specificity and efficiency of the N probesets.

### 2.4. Nanostring workflow

Comprehensive details of probes (consisting of sets of a fluorescently barcoded reporter probe paired with a capture probe, representing the Nanostring nCounter Elements chemistry) designed and used in this study have been provided in **Supplementary Table 1** and rationale for their choice is detailed in Section 3.1. The Nanostring nCounter Elements kit includes six External RNA Controls Consortium (ERCC) synthetic oligos with no known biological sequence homology, used as negative controls and to establish background signal for each run. The kit also includes six positive control DNA targets, representing a linear increase in concentration, which are employed together with their complementary probes to determine hybridization efficiency, linearity, and dynamic range of each run. Probes were chosen to target the same open reading frame (ORF) as the associated PCR assay where possible, while also considering the additional thermodynamic constraints of NNS probes. This process was done in consultation with Nanostring Technologies application specialists, resulting in two or three probes synthesized for each target. The exception was the single *Borrelia spp.* 16S rRNA gene probe. All probes were confirmed to be specific for their intended targets *in silico* using NCBI-BLAST.

For quantifying RNA targets, a 5 nM master stock of “core” reporter probeset (“A”) was prepared by diluting 5 μL of each 1 μM reporter probe in a total volume of 1 mL of TE buffer (Invitrogen). A 25 nM master stock of capture probeset (“B”) was prepared in parallel by diluting 5 μL of each 5 μM capture probe in a total volume of 1 mL of TE buffer (Invitrogen). Probes A and B working pool dilutions were then prepared separately by diluting 4 μL of each master stock with 29 μL of TE-Tween (10 mM Tris pH 8, 1 mM EDTA, 0.1% Tween-20). The hybridization master mix was prepared by combining 70 μL of hybridization buffer (Nanostring), 7 μL each of probe A and B working pools, and 28 μL of core nCounter XT TagSet (Nanostring). Hybridization was then carried out with 8 μL of the hybridization master mix and 7 μL of template RNA in a thermocycler at 67° C for 18 h (Bio-Rad Laboratories, Richmond, CA). The template RNA used for hybridization was either directly extracted using the RNeasy PowerFecal kit or, in addition, concentrated using RNA Clean & Concentrator-5 kit (Zymogen) or alternatively pre-amplified RNA using the Nanostring low RNA input kit as per the manufacturer’s instructions (c.f., Discussion).

For quantification of DNA targets, master stocks of “extension” reporter (“A”) and capture (“B”) probesets were prepared in the same manner as for the RNA core probesets. Similarly, working pools of extension probe A and B were created by diluting 4 μL of master stocks of extension tagset probe A and B with 29 μL of TE-Tween. A hybridization master mix containing both core and extension probesets was assembled according to the manufacturer’s instructions and contained 70 μL of hybridization buffer, 7 μL each of core tagset working pools probe A and B, 7 μL each of extension tagset working pools probe A and B, 28 μL each of core and extension TagSet reagents. Hybridization was carried out with 11 μL of this hybridization master mix and 4 μL of fragmented, denatured DNA for 18 h at 67 °C using a thermocycler (Bio-Rad Laboratories, Richmond, CA). 25 μL of extracted DNA was used to obtain 50 μL of fragmented DNA using the AluI endonuclease restriction enzyme (Biolabs, USA) as per the manufacturer’s manual. Fragmented DNA was denatured by heating at 95 °C for 5 min and immediately placed on ice before hybridization.

Post-hybridization, 20 μL of DNAse/RNase free water was added to the hybridized mix and ∼35 μL of the mix was loaded onto a single sample lane of a nCounter Sprint microfluidics cartridge (Nanostring) within 15 minutes. Each microfluidic cartridge has 12 lanes. All samples were run as technical duplicates in two lanes of the same microfluidics cartridge. A non-template control (NTC) comprising of DNAse/RNase free water used in the post-hybridization step was included along with five different sludge samples when testing for DNA targets to rule out any cross-contamination during the loading of samples into the cartridge. Furthermore, NTC and the five nucleic acid extracts were re-tested so that a total of 3 independent runs were achieved (3 cartridges) to assess variation within and between cartridges/runs.

Data was collected using either nCounterMAX or nCounter Sprint system and analyzed with the nSolver 4.0 software (Nanostrings). Six synthetic positive controls of known concentration and six synthetic negative controls were included in the Nanostring assay. As a quality control (QC) measure, assay linearity (R^2^) plotted between number positive counts and known concentration of each sample should be greater than 0.95, suggesting successful hybridization and pipetting accuracy. Once the QC criteria was met, the concentration of all targets were normalized as per Nanostring’s guidelines (Nanostring Technologies, 2017) and reported as normalized nanostring counts (nc). Briefly, the geometric mean of the positive controls for each sample was calculated, followed by the calculation of arithmetic mean of all geometric means of 12 samples. Individual normalization factor for each sample was then calculated by dividing the arithmetic mean by the geometric mean of that sample. Finally, the number of counts of each target gene was then multiplied with sample-specific normalization factor to obtain the normalized nc. Normalized counts from duplicate lanes were averaged creating 15 datapoints for the DNA targets (5 extracts repeated in 3 independent runs).

### 2.5. ddPCR workflow

All ddPCR runs were performed in 20 μL reactions mixtures using the QX200 droplet digital PCR system (Bio-Rad). For quantification of RNA targets, 1-Step RT-ddPCR Advanced Kit for Probes (Bio-Rad) was used and reaction mixture composed of 5 μL Super mix, 2ul of Reverse transcriptase, 1 μL of 300mM DTT, 1 μL of template RNA, and primers and probe at a final concentration of 900 and 250 nM, respectively. Similarly, ddPCR Supermix for Probes (Bio-Rad) was used to quantify DNA targets and reaction mixture composed of 10 μL of 2X ddPCR Supermix for probes, 900 nM primers and 250 nM probe, and 1 μL of AluI-digested DNA). The complete list of primers and probes used in the study is given in supplementary file (**Supplementary Table 2**). All RNA targets were tested in triplicate, while all DNA targets were tested in duplicate. The number of droplets formed ranged from 11358 to 19418.

### 2.6 Statistical analysis

The equation used to calculate the limit of detection (LOD) of NNS is given in Equation 1 (Crossland et al., 2023). Coefficient of variance (CV) was calculated as the ratio of standard deviation to the average. Spearman’s rank correlation test and Fisher’s exact test were performed to compare the results obtained using ddPCR and NNS platforms. Significance level was set at a *p* value of < 0.05. Data curation, analysis, and visualization were performed using Microsoft Office Excel 2019 (Microsoft Corporation, Redmond, USA) and GraphPad Prism 9 (GraphPad Software, San Diego, USA).

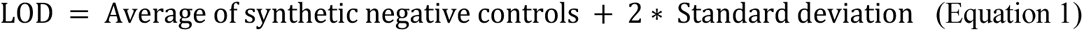

## 3. Results

### 3.1. Quantification of DNA targets in primary sludge samples

The LOD of our assay was set at 4.8 normalized counts based on the non-targeting controls (nc; see Materials and Methods). Specificity of the probes was assessed with five sludge-derived DNA extracts. For non-targeting probes, or for probes that target an RNA biomarker such as PMMoV (this RNA virus does not produce DNA intermediates), or luciferase (which can exist as either DNA or RNA but not expected to be present in wastewater) we observed an average nc close to 1 and never exceeding the LOD (**Fig. 1A**). The background counts with the PMMoV probe also indicate that the extract was free of RNA, as PMMoV RNA was present in high amounts in RNA extracts (cf. **Fig. 3A**). The dynamic range and accuracy of the instrument can be demonstrated by observing the nc of the spike-in positive controls that span four orders of magnitude from 125 aM to 128 fM (**Fig. 1A**; note the 128 fM counts are not shown to enhance clarity). Two No-template control (NTC) lanes were included in each run where water was used in place of wastewater extract to gauge cross-contamination of lanes. None of the DNA targets, except bacterial 16S rRNA genes were detected more than once in NTC lanes above LOD (**Fig. 1A**, violin plot). The counts of 16S in NTC lanes were close to LOD and likely indicate present, but negligible contamination of reagents or the water with bacterial DNA. The work environment routinely employs *E. coli* in molecular biology work, and this could be a source of contamination. Counts associated with the 16S #1 probe (16S-1) and *bla*_CMY_ #2 probe (*bla*_CMY_-2) were rejected from final analysis because they were found to contain sites for cleavage by the AluI enzyme which we used to fragment genomic and plasmid DNA to allow for more efficient hybridization.

We binned each probe for DNA targets into low, medium, high nc, and present these in **Fig. 1B-D**. We did not observe MPXV or Borrelia nc above LOD. The vancomycin resistance marker (vanA) nc was below LOD for all sludge samples, except for one sample replicate. Vancomycin resistance is still uncommon in Canada (0.36 per 10,000 patient days across all hospitals in 2022 (Public Health Agency of Canada, 2022). *bla*_OXA-48_ nc was detected more often and in more of the collected sludge samples. Among all the DNA targets tested, *bla*_CTX_, *bla*_KPC_, *bla*_CMY_, 16S, and crAssphage, were robustly detected in every sample in all 3 replicates (**Fig. 1C and 1D**). The nc of *bla*_CTX_ *bla*_OXA-48_, *bla*_KPC_, *bla*_CMY_, were found to be approximately one order of magnitude lower than that of 16S, crAssphage, ranging from 5.4–8.6, 25.4–152.7, 25.2–76.2, 33.9–92.3, 545.3– 2450.9, 836.6–1545.1 nc, respectively. The average intra- and inter-cartridge CV% among the replicates when quantifying 16S and crAssphage were 5.5 ± 6.8 and 9.3 ± 5.9, 12.4 ± 6.1 and 23.0 ± 3.6, respectively, indicating reproducibility and repeatability of the results.

To benchmark the accuracy of our NNS findings, we performed ddPCR on the same DNA extracts with published assays. ddPCR detected and quantified the low-copy targets *bla*_OXA-48_ and *vanA* in replicates of all wastewater samples (2.2-10.8 copies/μl) along with the medium- and high-copy targets (**Table 1**). In contrast, *bla*_OXA-48_ and *vanA* were detected in 4 out of 5 samples and 1 out of 5 samples, respectively by NNS. However, their detection could not always be replicated across multiple runs. A significantly lower positive ratio for *vanA* was observed when analyzed using NNS compared to dPCR (Fisher’s exact test, p < 0.05). This indicates that NNS has lower sensitivity compared to dPCR. ddPCR mirrored the quantitative results obtained by direct detection using NNS for medium- and high-copy targets. The approximate threshold detection for DNA targets appeared to be approx. 10 ddPCR-equivalent copies/μl of extract. Although this number may be sample-dependent with possible matrix effects on probe hybridization efficiency and/or enzyme-dependent PCR. Overall, these results highlight the specificity and sensitivity of NNS to directly detect multiple DNA targets of interest in a single extract from wastewater.

**Table 1.** Number of replicates and concentration of ARGs and 16S DNA in primary sludge samples.

| Sample ID | DNA<br>conc (µg/<br>µL) | No. of positive/no. of replicates tested (Concentration*) |  |  |  |  |  |  |  |  |  |  |  |  |  |
| --- | --- | --- | --- | --- | --- | --- | --- | --- | --- | --- | --- | --- | --- | --- | --- |
|  |  | ddPCR (copies/µL) |  |  |  |  | NNS (normalized counts) |  |  |  |  |  |  |  |  |
|  |  | *bla*<sub>OXA-48</sub> | *vanA* | *bla*<sub>KPC</sub> | *bla*<sub>CTX-M</sub> | 16S | *bla*<sub>OXA-48</sub><br>A | *bla*<sub>OXA-48</sub><br>B | *vanA* A | *vanA* B | *bla*<sub>KPC</sub> A | *bla*<sub>KPC</sub> B | *bla*<sub>CTX-M</sub><br>A | *bla*<sub>CTX-M</sub><br>B | 16S |
| PS-1 | 495 | 2/2 (8.2) | 2/2 (3.3) | 2/2 (2025.5) | 2/2 (926.5) | 2/2 (3.1 × 10<sup>4</sup>) | 0/3 (N/A) | 1/3 (5.9) | 0/3 (N/A) | 1/3 (8.2) | 3/3 (67.2 ± 19.2) | 3/3 (36.2 ± 8.1) | 3/3 (45.0 ± 20.8) | 3/3 (53.6 ± 20.8) | 3/3 (1144.5 ± 110.5) |
| PS-2 | 745 | 2/2 (9.2) | 2/2 (3.5) | 2/2 (2736) | 2/2 (1112) | 2/2 (4.1 × 10<sup>4</sup>) | 1/3 (7.8) | 1/3 (7.3) | 0/3 (N/A) | 0/3 (N/A) | 3/3 (125.7 ± 37.7) | 3/3 (48.3 ± 7.2) | 3/3 (42.3 ± 10.5) | 3/3 (64.4 ± 16.3) | 3/3 (1349.6 ± 161.7) |
| PS-3 | 572 | 2/2 (7.1) | 2/2 (2.2) | 2/2 (1897) | 2/2 (907) | 2/2 (2.9 × 10<sup>4</sup>) | 2/3 (6.5 ± 0.1) | 1/3 (5.4) | 0/3 (N/A) | 0/3 (N/A) | 3/3 (72.3 ± 10.7) | 3/3 (29.9 ± 5.0) | 3/3 (33.2 ± 8.8) | 3/3 (50.8 ± 15.7) | 3/3 (1040.9 ± 86.8) |
| PS-4 | 972 | 2/2 (10.8) | 2/2 (3.9) | 2/2 (3658.5) | 2/2 (1695) | 2/2 (4.2 × 10<sup>4</sup>) | 2/3 (7.5 ± 1.6) | 2/3 (6.5 ± 0.0) | 0/3 (N/A) | 0/3 (N/A) | 3/3 (126.3 ± 28.0) | 3/3 (49.2 ± 13.2) | 3/3 (55.8 ± 14.4) | 3/3 (90.8 ± 22.0) | 3/3 (1078.1 ± 247.9) |
| PS-5 | 523 | 2/2 (7.1) | 2/2 (3.6) | 2/2 (2867) | 2/2 (1088) | 2/2 (2.0 × 10<sup>4</sup>) | 2/3 (6.8 ± 1.4) | 0/3 (N/A) | 0/3 (N/A) | 0/3 (N/A) | 3/3 (107.8 ± 26.3) | 3/3 (41.3 ± 9.3) | 3/3 (39.3 ± 20.4) | 3/3 (58.2 ± 20.4) | 3/3 (952.7 ± 86.7) |

\*Average ± Standard deviation; Two replicates represent 2 wells for ddPCR and 3 replicates represent results summarized from 3 cartridges.

### 3.2. Direct detection of in vitro transcribed SARS-CoV-2 N RNA

The N probesets of SARS-CoV-2 for NNS were designed to be overlapping with or proximal to the USCDC N1 and N2 PCR loci, as they have consistently provided the highest analytical sensitivity and are widely used in wastewater applications (Ahmed et al., 2022, Perez-Cataluna et al., 2021) (**Fig. 2A**). Critically, the normalized RNA counts directly determined using NNS and specific to the multiplexed N ORF probesets (COV2N1, COV2N2, COV2N3, COVN4) displayed high linearity (R2 ≥ 94%) across the copy number range and correlated well with copy numbers determined in singleplex N1 RT-ddPCR reactions (**Fig. 1A**). The multiplexed nature of NNS allowed us to simultaneously assess specificity by looking at the counts obtained from off-target (i.e., not in N RNA). Indeed, both PMMoV probes (PMMOV_1 and PMMOV_2) (**Fig. 2C**) were below the LOD in all titrations of the SARS-CoV2 N RNA, indicating high specificity of NNS.

**Figure 2.**
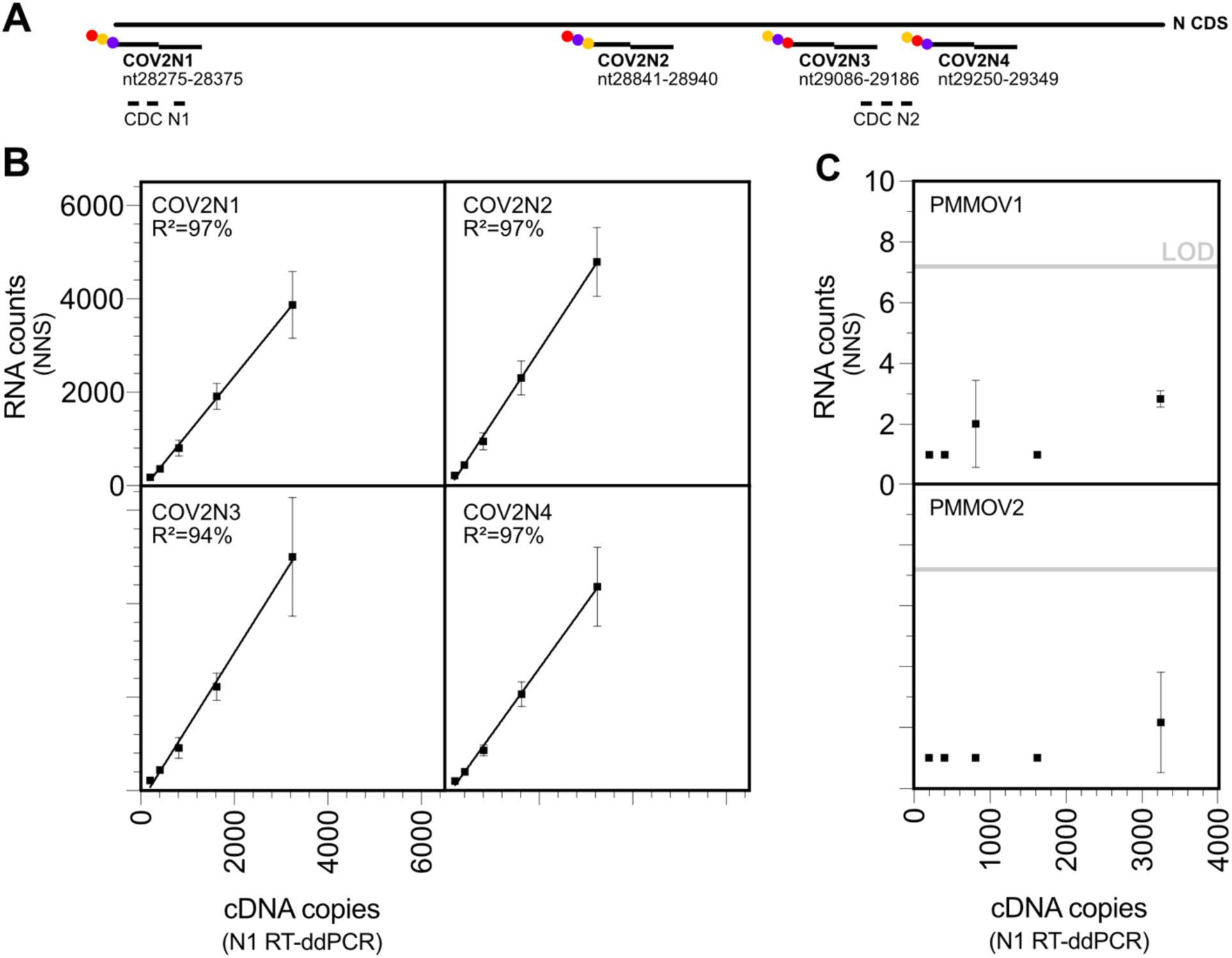
NNS vs. RT-ddPCR performance on in-vitro transcribed SARS-CoV-2 N ORF RNA template. (A) The four NNS bi-partite probes (reporter and capture probes) shown were designed to target the SARS-COV-2 N (Wuhan) ORF with a preference to be close to either the CDC N1 or CDC N2 PCR loci. (B) Correlation plots showing the linear relationship between multiplexed, direct molecule counting by NNS at four different genetic loci and proxy counting of amplified cDNA by RT-ddPCR using the singleplex N1 assay. Lines of regression are plotted and fit of the line (R^2^) indicated for each of the four probes. (C) In the same lanes, specificity was demonstrated with counts below the LOD for probes targeting PMMoV RNA.

### 3.3. Direct detection and multiplexed quantification of PMMoV and SARS-CoV-2 RNA in primary sludge samples

Five frozen and archived primary sludge samples (approx. 1.5 years old at the time of extraction) representing a 6-week period in spring of 2022 during the tail end of the BA.2 epi-wave were thawed and the RNA extracted. Based on preliminary experiments and to maximize sensitivity we chose to further concentrate 100 μl RNA extracts down to 15 ul (cf. Materials and Methods) and use the nCounter MAX instrument instead of the nCounter Sprint that we used to detect the AMR biomarkers. The nCounter MAX uses the same assay chemistry but has the advantage of less dead volume (100% vs. 30% of the hybridized material is loaded) and approximately twice the number of imaged fields that combined to effectively lower the LOD in our hands (data not shown). We were able to directly detect and quantify PMMoV RNA with the two PMMoV-specific probes in all five of the tested primary sludge extracts. RT-ddPCR targeting PMMoV confirmed its presence in concentrations ranging from 3300-5900 RNA copies/ul in the concentrated extract. Demonstrating the accuracy of NNS, a strong linear correlation between RNA counts and RT-ddPCR-generated copies was observed (r ≥ 0.9, *p* < 0.05) with strong linearity (R^2^>0.80) observed across samples (**Fig. 3**). Notably, there was not a 1:1 relationship observed for counts and copies, which argues for a need for calibration in WWS applications. An effective assay requires that the variability of technical replicates is minimal. We compared the percent coefficient of variation (%CV) of two technical replicates between platforms; our two NNS PMMoV probes (two sample lanes on the same chip/run) with gold standard RT-qPCR and RT-ddPCR PMMoV assays (two wells on the same plate/run). **Fig. 3B** shows the %CV of these technical replicates across the five sludge extracts used in **Fig. 3A** for NNS and RT-ddPCR and four different sludge extracts for RT-qPCR, all performed by the same operator. Together, these data show that direct, enzyme-independent detection of PMMoV RNA with NNS performs similarly to NAATs that depend on indirect, proxy measurement of amplified DNA. For this reason, we considered the average NNS signal from both PMMOV_1 and PMMOV_2 probes in further analysis below.

**Figure 3.**
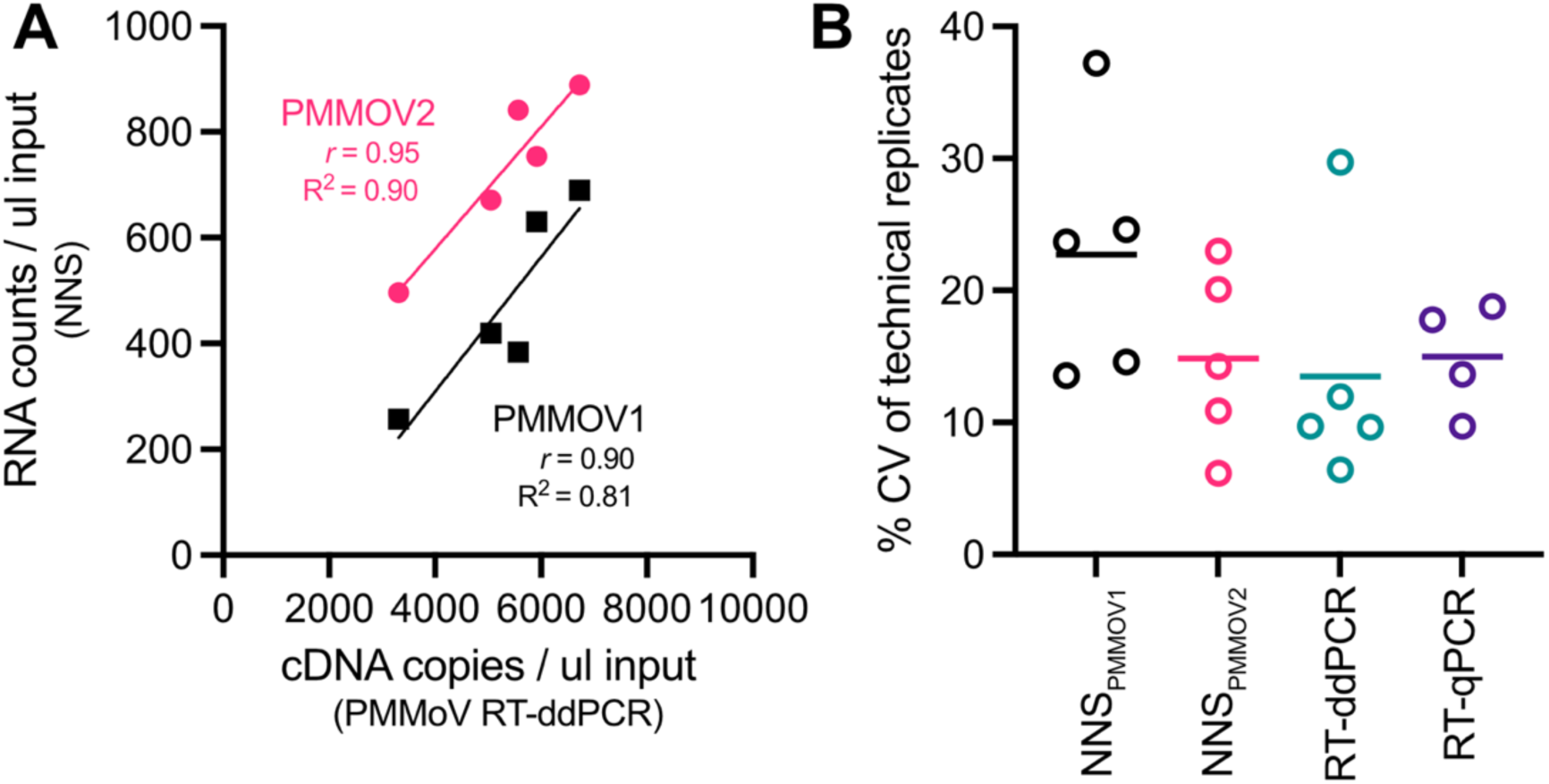
Accuracy of direct and multiplexed PMMoV detection and quantitation in primary sludge. (A) Correlation and linear relationship of NNS PMMOV_1 and PMMOV_2 probe counts with PMMoV RT-ddPCR across five different primary sludge samples expressed per ul of extract assayed. Each datapoint represents the mean of counts for two NNS lanes and four RT-ddPCR wells (B) Percent CV of RNA extract replicates across five samples for NNS and RT-ddPCR or four other samples for RT-qPCR.

We assessed the ability of NNS to follow trends in SARS-CoV-2 RNA levels in wastewater over time. For this we examined the SARS-CoV-2 specific N signal from the NNS experiment performed in the previous section which represents a waning outbreak of COVID-19 across 6 weeks. Our four N probes were able to reliably detect SARS-CoV-2 N RNA from the re-extractions of archived wastewater samples that was above the signal observed for no-template controls (**Fig. 4A**, left and right plots, respectively). Specificity was demonstrated with the average counts from 6 off-target oligonucleotides (“NEG” in **Fig. 4A**) mirroring NTC counts. There was a clear difference in sensitivity between probes although they all were closely correlated over time. Probe N_1 consistently demonstrated higher counts relative to N_2, N_3, N_4 across all trials, although it’s important to note that N_1 also consistently showed higher background in NTC lanes.

**Figure 4.**
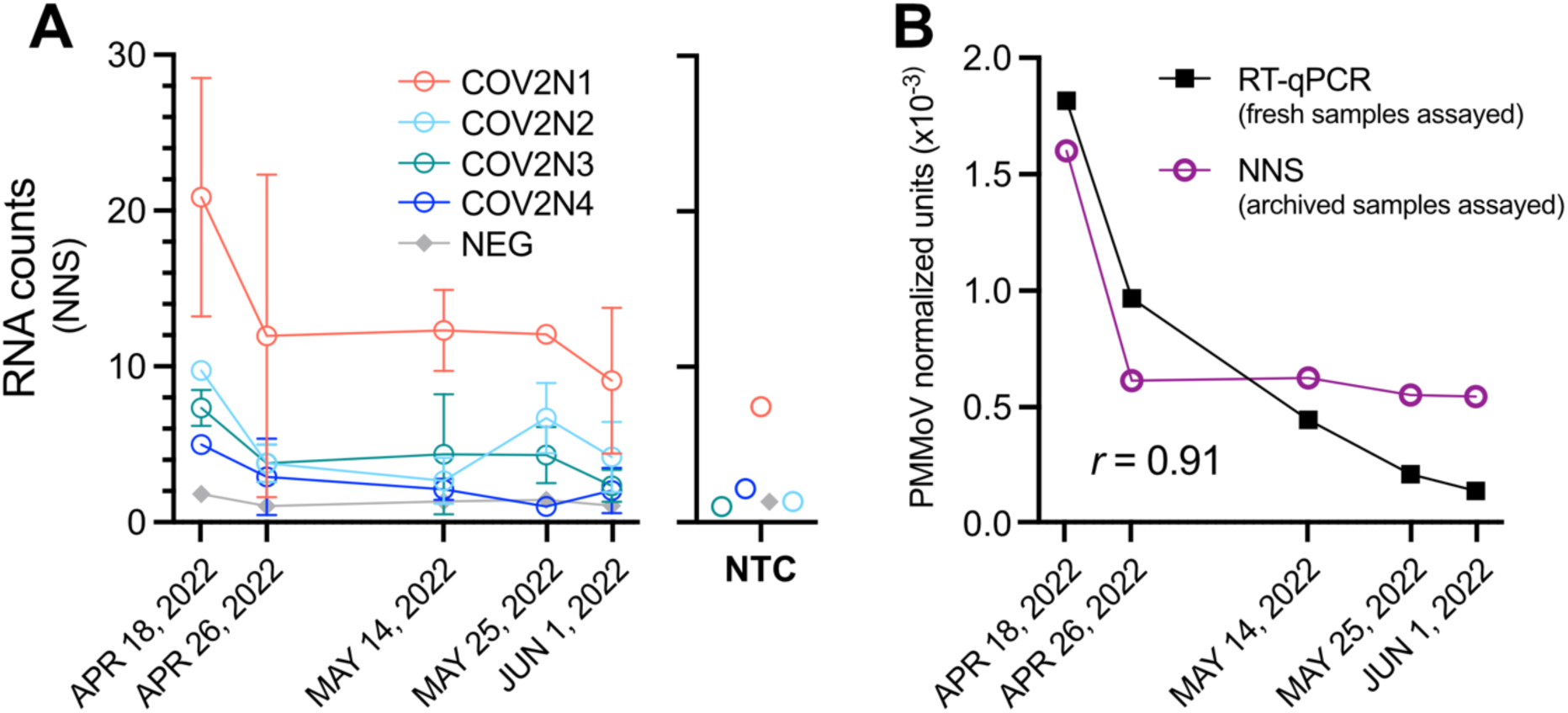
Direct quantification of SARS-CoV-2 and PMMoV RNA in wastewater by NNS in the waning period of a COVID-19 outbreak. (A) Mean NNS Counts using the 4 N RNA probes. Error bars represent s.d. of two technical replicates. The average counts associated with six oligonucleotide probes that do not target known RNA sequences (NEG) were close to 1 (the minimum counts possible) demonstrating specificity of the N probes. Similar counts were found in no template control lanes (NTC; plot on right showing mean of 2 replicates for each probe) apart from probe COV2N1 which exhibited higher counts in NTC as well as wastewater extracts. (B) The mean of counts of all four N probes in (A) was normalized to the mean of both PMMoV probes used in Fig. 3A and this measure was compared to the equivalent RT-qPCR-derived metric (N1+N2/PMMoV) that was derived from fresh samples approx. 1.5 years prior to the NNS experiment with archived samples. Pearson’s correlation of the two metrics is shown.

Like many WWS programs worldwide, the Wastewater Surveillance Initiative in Ontario, Canada has used RT-qPCR to quantify SARS-CoV-2 RNA based on the average combined signal of the USCDC N1 and N2 assays (Thakali et al., 2024). This metric of SARS-CoV-2 RNA concentration is routinely normalized to that of PMMoV, thus controlling for variation in contributing human excreta; then reported to public health agencies. These values were derived from fresh samples at the time of collection, and we used these values to compare with our NNS-derived equivalent metric on the designated sample dates. As shown in **Fig. 4B**, a decrease in PMMoV-normalized SARS-CoV-2 RNA signal in wastewater was observed from April 18, 2022 to June 1, 2022 using NNS. A similar decreasing trend was observed using the RT-qPCR-equivalent metric derived from fresh samples; however, a plateau phase was noted in the trend generated by NNS, suggesting a lower sensitivity of NNS compared to qPCR (**Fig. 4B**). Correlation analysis between PMMoV-normalized concentrations measured by qPCR and NNS revealed significant association (r = 0.91, *p* < 0.05) (**Fig. 4B**). Together, these data indicate that NNS could be used to accurately follow quantitative trends in low-copy RNA targets such as SARS-CoV-2, although analytical sensitivity is lower than platforms based on nucleic acid amplification.

## 4. Discussion

WWS holds great potential to monitor a broad range of nucleic acid-based biomarkers. Tools with multiplexing capabilities that facilitate their tracking need to be explored. In this study, we have applied NNS for measuring diverse DNA and RNA targets of public health significance in nucleic acid extracts of primary sludge collected from a WWTP. As a pilot study, we optimization nucleic acid extraction and concentration, and ran two parallel hybridization workflows for DNA and RNA targets to maximize sensitivity. Contaminating DNA or RNA can lead to lower counts due to reduced hybridization efficiency and a lower effective DNA input amount caused by the overestimation of DNA concentration when measured by UV absorbance (Nanostring Technologies, 2020). Therefore, RNA was removed with RNase despite using a kit that facilitates extraction of both DNA and RNA. Complete removal of RNA by our protocol was verified by the failure to detect PMMoV by RT-qPCR (data not shown). Nanostring Technologies recommends fragmenting DNA by AluI digestion prior to hybridization to prevent binding hindrances with probes, as the higher order structures nucleated by long stretches of genomic DNA cause steric hindrance of probe binding relative to shorter RNA fragments which are comparatively easier to denature at the hybridization temperature employed in the process. DNA fragmentation post-digestion was also confirmed by visualization via gel electrophoresis (data not shown). One disadvantage of DNA digestion using AluI is the possibility of inaccurate or failed hybridization if the genomic region targeted by the NNS probe contains an AluI sequence. Ultrasonication is an alternative method for DNA fragmentation in these cases (Tolman et al., 2024).

To rule out false positive and negative counts, a threshold value or Limit of Detection (LOD) was calculated as per the guidelines of Nanostring Technologies (Equation 1). Recently, three levels of LOD stringency—low, medium, and high—have been recommended for robust data quality control, enabling researchers to choose the level of stringency most suitable for their datasets (Crossland et al., 2023). The medium LOD corresponds to the manufacturer’s LOD guideline, whereas the low LOD, calculated as the mean of all synthetic negatives (i.e., non-targeting probes), serves as a less conservative threshold (Crossland et al., 2023). Similarly, the high LOD, calculated as twice the medium LOD, can be used to eliminate rare false positives (Crossland et al., 2023). Reproducible quantification is a pre-requisite for any platforms used in research or diagnostics and this study highlights the reproducibility of the NNS based on the concentration of crAssphage and 16S DNA markers. It should be noted that non-nc of *bla*_CTX_, *bla*_KPC_, and *bla*_CMY_ were within the same range as those of synthetic positive controls E (0.5 fM) (12-135 non-nc) and F (0.125 fM) (3-70 non-nc). This indicates that the concentration of *bla*_KPC_ and *bla*_CTX-M_ were near threshold level of quantitation and thus precision may be affected.

Despite optimized sample processing, this study shows that NNS has a lower sensitivity compared to gold-standard NAATs that are widely used in WWS applications. Nanostring technologies recommends 150 ng of DNA and 100 ng of RNA for all NNS assays. The lower sensitivity can be explained by the probable low concentration of DNA/RNA marker in wastewater. Precipitation of nucleic acids with ethanol (Green and Sambrook, 2020) after nucleic acid extraction using commercial kits can be a cost-effective method to further concentrate the extracted nucleic acids and future studies on modifying protocol to improve sensitivity of NNS is encouraged. Despite the lower sensitivity, a potential advantage of NNS is that it doesn’t rely on enzymatic reactions. This suggests that samples exhibiting PCR inhibition may be less susceptible to test failure using NNS. Importantly, pre-amplification could also be used to increase the sensitivity of NNS (i.e., for RNA targets, RT followed by several cycles of PCR or only the latter for DNA targets). This would increase the number of quantifiable low copy targets but would make the platform enzyme dependent. Notably, it might also be possible to directly quantify high-copy targets (such as fecal content markers) from raw or filtered wastewater or other environmental sample types with minimal processing, and this remains to be investigated. During the COVID-19 pandemic, WWS has been greatly utilized to determine the circulating variants of SARS-CoV-2 via sequencing. In general, a sample with a qPCR cycle threshold value of 37 or lower with the US CDC N1 assay is recommended for sequencing. Our experience indicates that any sample with NNS counts above the High LOD is likely suitable for sequencing due to lower sensitivity and the rarity of false-positive results when using the NNS.

As a limitation, this pilot study did not directly assess the need to calibrate NNS counts with those derived by dPCR. However, normalizing to a PMMoV marker (or equivalent) as we show in **Fig. 4B** is an effective calibration method. A similar approach could be used to report AMR counts, perhaps using crAssphage and/or 16S DNA counts (**Fig. 1B**), although further validation would be needed. We also acknowledge that the requirement of overnight hybridization (16+ hours), potentially reduces any perceived benefit as an early warning system, although the reduced hands-on time and ease of interpretation as compared to NAATs should be considered. We did not perform an in-depth cost analysis, however the cost-advantage of multiplexed NNS would quickly become apparent compared to singleplex dPCR with panels of more than 10 target probes.

Nevertheless, the multiplexable nature of NNS opens the door to quantifying hundreds of targets from a single nucleic acid extract. It can be easily adapted to target multiple pathogens by simply adding probes into the extension tagsets or creating a larger custom core tagset. It is also conceivable that nucleic acid-tagged antibodies or aptamers could be used to quantify protein or small molecule targets with the platform. NNS is economical compared to dPCR but more expensive compared to qPCR when only testing for a single or limited number of targets. However, when multiple targets are screened using NNS, the cost and hands-on time for sample preparation is significantly reduced. Therefore, NNS has the potential be a valuable tool for initial screenings and as part of a broader wastewater monitoring strategy in communities.

## 5. Conclusion

This study successfully demonstrated the application of NNS to quantify targets of public health significance. Due to the lower sensitivity of NNS compared to NAATs and the requirement of an overnight hybridization step, NNS may not be suitable to provide early warning via WWS. However, as the scope of WWS expands to monitor a broad range of nucleic acid-based targets, the multiplexing capability of NNS (i.e., providing for economy of scale on a per-target basis), combined with reduced hands-on time, suggests that it has significant potential in WWS.

## Data Availability

All data produced in the present study are available upon reasonable request to the authors.

## CRediT authorship contribution statement

Ocean Thakali: Methodology, Formal analysis, Investigation, Writing – original draft, Walaa Eid: Methodology, Formal analysis, Investigation. Julia Brasset-Gormy: Methodology, Investigation. Sean Stephenson: Investigation. Tyson Graber: Supervision, Funding Acquisition, Conceptualization, Writing – review, editing, and finalization. All authors reviewed the manuscript.

## Acknowledgements

We thank Dr. Katey Rayner at the University of Ottawa Heart Institute for providing access to the Nanostring nCounter MAX. We are grateful to Jade Daigle and Chand Mangat who were instrumental in speeding up this project by providing a short list of ARGs to target in wastewater and associated qPCR assays based on work from her master’s thesis.

## SUPPLEMENTARY MATERIAL

**Table S1.** Nanostring probes used in this study (XLSX file).

**Table S2.** ddPCR primers and probes used in this study (XLSX file).

